# Algorithm for Predicting Valvular Heart Disease from Heart Sounds in an Unselected Cohort

**DOI:** 10.1101/2022.11.28.22279153

**Authors:** Per Niklas Waaler, Hasse Melbye, Henrik Schirmer, Markus Kreutzer Johnsen, Tom Dønnem, Johan Ravn, Stian Andersen, Anne Herefoss Davidsen, Juan Carlos Aviles-Solis, Michael Stylidis, Lars Ailo Bongo

**Affiliations:** Department of Computer Science, UiT The Arctic University of Norway, Norway; General Practice Research Unit, Department of Community Medicine, UIT The Arctic University of Norway, Norway; Department of Cardiology, Akershus University Hospital, Norway; Institute of Clinical Medicine, Cardiovascular Research Group, University of Oslo, Norway; Department of Clinical Medicine, UiT The Arctic University of Norway, Norway; Medsensio AS, Norway; Levanger Hospital, Nord-Trøndelag Health Trust, Norway; Sørbyen Legesenter, Tromsø, Troms og Finnmark, Norway

## Abstract

**Background:** Although neural networks have shown promise in classifying pathological heart sounds (HS), algorithms have so far either been trained or tested on selected cohorts which can result in selection bias. Herein, the main objective is to explore the ability of neural networks to predict valvular heart disease (VHD) from recordings in an unselected cohort.

**Methods and results:** Using annotated HSs and echocardiogram data from 2124 subjects from the Tromsø 7 study, we trained a recurrent neural network to predict murmur grade, which was subsequently used to predict VHD. Presence of aortic stenosis (AS) was detected with sensitivity 90.9%, specificity 94.5%, and area-under-the-curve (AUC) 0.979 (CI:0.963-0.995). At least moderate AS was detected with AUC 0.993 (CI:0.989-0.997). Moderate or greater aortic and mitral regurgitation (AR and MR) were predicted with AUC 0.634 (CI:0.565-703) and 0.549 (CI:0.506-0.593) respectively, which increased to 0.766 and 0.677 when adding clinical variables as predictors.

Excluding asymptomatic cases from the positive class increased sensitivity to AR from 54.9% to 85.7%, and sensitivity to MR from 55.6% to 83.3%. Screening jointly for at least moderate regurgitation or presence of stenosis resulted in detection of 54.1% of positive cases, 60.5% of negative cases, 97.7% of AS cases (n=44), and all 12 MS cases.

**Conclusions:** Despite the cohort being unselected, the algorithm detected AS with performance exceeding performance achieved in similar studies based on selected cohorts. Detection of AR and MR based on HS audio was unreliable, but sensitivity was considerably higher for symptomatic cases, and inclusion of clinical variables improved prediction significantly.

## INTRODUCTION

Valvular heart disease (VHD) is a major source of dysfunction, reduced quality of life, early death and increased health-care costs^1–3^. The prevalence of VHD in the USA is estimated to be 2.2% in the general population, and 13.3% amongst those 75 or older^4^. The prevalence of aortic stenosis (AS), the most common form of VHD in developed countries with a poor prognosis when left untreated, was estimated to 12.4% in those 75 years or older based on data from European countries and North America in 2013^5^. Prevalences of mitral regurgitation (MR) and aortic regurgitation (AR) are also strongly age dependent^6,7^. With the projected growth of the elderly population, and the rapid increase of VHD prevalence with age^8^, the societal burdens associated with VHD are expected to increase considerably^2^.

Although studies have indicated that the stethoscope can be a cost-effective screening-tool for cardiovascular disease, its potential utility is limited by the increasing time constraints imposed on doctors, the declining skill of health care providers in performing cardiac auscultation^9–11^, and low interrater agreement. While the echocardiogram is accurate, it is expensive^12^ and requires highly trained personnel to analyze its results, and therefore can not replace the stethoscope as a front-line screening tool, especially in low-income regions where its availability is even more limited. Effective treatment of VHD exists^13^, and evidence suggests that earlier detection might be associated with better outcomes^14^. These facts, together with the high prevalence of undiagnosed VHD in the elderly population^15^, stresses the need for inexpensive and widely available screening methods. Point-of-care ultra sound has been launched as a potential solution. However, it will take many years before most general practitioners (GPs) are fully trained in this, and it has not been clearly demonstrated to improve upon auscultation in detecting AS^16–18^. Adding this to the physical examination will not help with the time constraints either.

Due to the aforementioned limitations of cardiac auscultation and echocardiography, and the success of machine learning methods in a wide range of classification problems, interest in automated HS analysis has increased considerably in recent years. Especially deep learning models have shown great potential for HS classification^19^, and the ability of algorithms to discriminate between normal and abnormal heart sounds has approached 100% accuracy in some studies^20^, although these results have yet to be validated on external datasets to demonstrate generalizability.

Automated HS-analysis could potentially offer several advantages over manually performed auscultation. It could make the procedure more widely available, which is especially important in low income regions where access to trained health-care personnel and echocardiogram may be severely limited. Increased availability could help reduce the high prevalence of undiagnosed VHD, and facilitate earlier detection of high risk individuals. Furthermore, automated HS-analysis could improve classification accuracy by reducing the influence of random or subjective factors, and it is not limited by human constraints such as limited range of hearing or time available for training.

Despite impressive results, the ability of state-of-the-art methods to effectively screen for undiagnosed VHD in a general population has not been demonstrated. Due to the rarity of significant VHD relative to the large amounts of data required to successfully train deep neural networks, preferentially recruiting sick individuals for study participation is common practice in data collecting data for machine learning studies. It is plausible that this practice results in high prevalence of cases that are more detectable and/or symptomatic, which in turn might bias test metrics^21^. This is a potential problem, since degree of detectability or presence of symptoms are not always reliable indicators of VHD severity^22–24^, and it is therefore important to assess the impact that the study participant selection procedure has on algorithm performance metrics. Our primary aim is to explore this effect and shed light on the potential of machine learning technology as a front-line screening tool for VHD in a general population. To assess this potential, we develop a murmur detection algorithm using state-of-the-art methods which we then use to screen for VHD. We also test whether the neural networks are able to extract additional diagnostic features beyond the presence of murmurs from the audio signal by training the network to predict VHD targets directly. We aim to establish which pathological cases are reliably detected, and how detectability relates to disease severity and presence of symptoms. Furthermore, we seek to establish if more accurate prediction of AS can be achieved by combining the murmur predictions of each of the four standard auscultation positions using a multivariate regression model. Finally, we aim to establish if clinical variables together with auscultation based algorithm predictions can improve VHD screening accuracy by using multivariate logistic regression models.

## METHODS

### Study Design

The Tromsø study is an ongoing population based prospective study that was initiated in 1974, in the municipality of Tromsø, Norway. The 7th and most recent survey was carried out in 2015 and 2016, which provided the data used in this study^25^. All inhabitants of Tromsø aged 40 or older (ages ranged from 40 to 99) were invited to participate in the study. The Tromsø study is the largest study of its kind in Norway, with 32 591 people invited to participate in its 7th iteration, of which 64.7% participated. A randomly selected subset of the participants underwent echocardiography examination (n=2340) and physical exams which included cardiac auscultation (n=2409) ^26^. In total, 2132 participants underwent both echocardiography and heart sound recording, and these constitute the dataset examined in this study. See **Figure 1** for a flow chart overview of the data collection.

**Figure 1.**
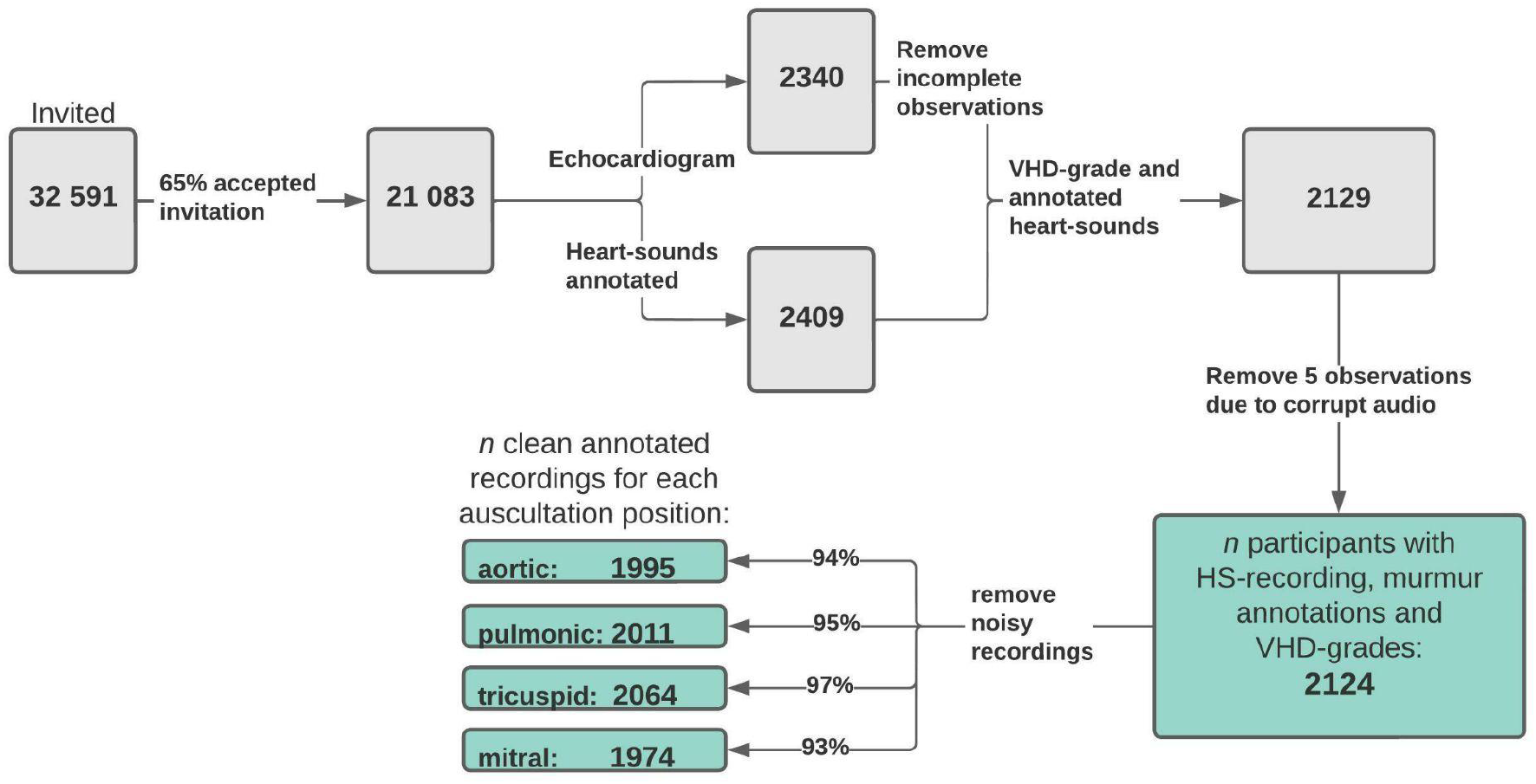
Data flow-chart. Flow chart overview of how the study dataset was formed, and how many samples were excluded due to noisy or incomplete data. The values at the end show the number of usable (absence of considerable noise) recordings annotated with murmur grade as well as grade of AR, MR, AS and MS for each auscultation position. VHD = valvular heart disease. AR = Aortic regurgitation. MR = Mitral regurgitation. AS = Aortic stenosis. MS = Mitral stenosis.

All study participants received questionnaires by email, and from the questionnaires we use information on the following variables: dyspnea, blood pressure, chest pain, angina pectoris, smoking status, and diabetes. For all participants, we have access to sex, age and body mass index (BMI), and from physical exams we have access to heart rate and pulse oximetry results (93.9% and 94.1%, respectively, of participants in our dataset have data on these variables). Proportions of missing data for the questionnaire variables are 4.24% (dyspnea), 2.45% (high blood-pressure), 1.7% (chest pain), 4.0% (angina), 1.4% (smoking status) and 2.8% (diabetes). More details on the clinical variables can be found in **Table S1**. In the multivariate analysis (see page 19 and 20), missing questionnaire answers were filled in using the most common category for the associated question. Given the low prevalence of missing values, we do not believe this to be a significant source of bias.

### Heart Sound Recording

In the Tromsø 7 study, a microphone was attached (Sennheiser MKE 2-EW) inside the tube of a stethoscope (Littmann classic II) 10 cm from the chestpiece. The microphone was connected to a wireless system (Sennheiser EW 112-P G3-G) that transmitted the sound signal to a computer via an external sound card (Scarlett 2i2, Focusrite Audio). Heart sounds were recorded for 10 seconds. Audio files were recorded in “.wav” format in a single monophonic channel with a depth of 16 bits at a rate of 44,100 hz. Participants were sitting in a chair and were asked to breathe normally. For each participant recordings were collected in four locations: aortic (2nd intercostal space, right parasternal line), pulmonic (2nd intercostal space, left parasternal line), tricuspid (4th intercostal space, left parasternal line), and mitral positions (5th intercostal space, left mid-clavicular line), which in the following will be referred to as positions 1-4 when convenient.

### Heart Sound Annotation

The heart sound recordings were annotated by two GPs (AD and SA) who were both working on PhD projects on heart sounds. AD is a GP specialist with two years training in cardiology. Ahead of the annotation, the two GPs, a professor of cardiology (HS), and a professor of General practice (HM) independently annotated 400 recordings, discussed all disagreements, and reached consensus on the presence and quality of murmurs. This training was reinforced at intervals throughout the annotation process when the two annotaters discussed disagreements and pursued consensus on the presence of murmurs. The consensus outcomes, to which HM and HS also contributed, are not dealt with in this paper. The annotators watched spectrogram visualizations of the recordings (using Adobe Audition CS6) while classifying the heart sounds. They were blind to the echocardiography results and other information about study participants during the HS annotation.

Each recording was annotated as either normal, systolic murmur, diastolic murmur, or noise (unable to annotate). Any perceived murmurs were graded on a scale from 1 (faint) to 6 (distinct), referencing the Levine scale, which is commonly used in clinical practice^27^. However, the scale is not directly transferable to recordings, as grades 4-6 are associated with a palpable thrill. Therefore, in our annotated set, grades higher than 3 only reflect increases in murmur loudness, and not other aspects associated with the levine scale. In total, 2129 participants were annotated with both murmur grade and VHD grade. Of these, 5 were removed due to corrupted audio-files, resulting in an effective sample of 2124 participants, and 8496 annotated audio files (with 1416 minutes of recorded audio). We trained the algorithm only on the recordings that both annotators had agreed were not noisy (recordings will hereafter simply be referred to as *noisy* and *non-noisy*), and attempted prediction only on non-noisy recording. In total, there were 129 (6.1%), 113 (5.3%), 60 (2.8%) and 150 (7.1%) noisy recordings in positions 1 to 4 respectively. 1.4% of participants had noise in all four recordings, and these observations were excluded from analysis related to algorithm predictions. The noisy recordings that were included were assigned a murmur grade of 0.

For algorithm training, we took the average of the murmur grade annotations to represent each position, and treated it as a continuous variable. In the following, this variable is referred to simply as *the* murmur grade. By this convention, the data-set contained a total of 465, 280, 303, and 196 cases of murmur grade>0 in positions 1-4 respectively, yielding a total of 1244 audio recordings for which at least one annotator perceived a murmur.

### Echocardiography

All echocardiogram examinations were performed according to the American Society of Echocardiography’s Guidelines using a GE Vivid E9 (GE Medical, Horten, Norway) ultrasound scanner^28^. The examination was performed by an experienced echo technician before the heart sounds were annotated, and reading of the results was performed by an experienced physician (MS)^26^. AS and MS severity was graded on a scale of 0 to 3 (absent, mild, moderate and severe), and AR and MR severity on a scale of 0 to 4 (absent, trace, mild, moderate and severe). AS was graded using the aortic valve mean pressure gradient (AVPGmean), using cutoff values 15 mm Hg (mild), 20 mm Hg (moderate) and 40 mm Hg (severe)^29^.

### Algorithm Development

The murmur detection algorithm was trained using HS recordings from all four auscultation positions, effectively treating the recordings of each individual as four independent observations. The model and data processing used in this study are based on a study by Latif et al., in which state of the art results were achieved for detecting abnormal heart sounds on the physionet 2016 challenge dataset^20^. For network architecture we used a 2-layer (50 neurons each) long-short-term-memory (LSTM) network, followed by a fully connected layer consisting of 30 neurons (**Figure S1)** shows a schematic overview of the model architecture). The network was trained to predict murmur grade as a continuous variable (the Supplementary Materials includes a performance comparison between models trained using binary and continuous labels respectively). The initial learning rate was set to 0.002, and was halved every 5 epochs. As the ratio of positive to negative cases was highly imbalanced, we resampled from the positive class (defined in this context as all observations with murmur grade≥ 1) until the ratio was approximately 1.

After performing spike removal (following the steps outlined in a 2009 paper by Schmidt et. al.^30^) and downsampling the signal to sampling rate of 2205 Hz, each recording was segmented into 6 overlapping (50%) blocks, each consisting of 4 cardiac cycles (with each cycle starting and ending at the first heart sound S1), using a modified version of the segmentation algorithm of Springer et al^31^. For each segment, the 13 first Mel frequency cepstral coefficients (MFCC) were computed, where we used the Hanning window function for computing the Fourier transform of the signal, with a time step size of 25 ms, and window overlap of 10 ms. The resulting MFCC matrix was then resized to dimensions 13×200 using cubic interpolation, forming the input units of data of the LSTM network. The predicted murmur grade of a recording was obtained by taking the median activation-value across the 6 extracted segments. **Figure S2** provides a schematic overview of the steps that convert raw audio input to algorithm prediction.

A frequently occurring error that we observed was that Springer’s segmentation algorithm sometimes failed to correctly estimate the heart rate despite the audio being of reasonably high quality. We therefore modified the publicly available code^32^ and implemented a method that utilizes all four HS recordings of an individual in order to produce a more robust heart rate estimate. Both modifications are described in detail in the Supplementary Materials.

### Predicting AS Using Multiple Auscultation Positions

We hypothesized that more accurate prediction could be achieved by aggregating murmur grade predictions from all four recordings in a linear multivariate predictive model, rather than predicting AS using audio from a single predetermined position. In the following, these models are referred to as the *multi-position model* and *single-position models* respectively. As there were only 45 cases of mild or greater AS in the development set, using AS classification accuracy for model selection would likely result in overfitting. Therefore, we first model AVPGmean using linear regression, and subsequently use the fitted model to predict AS (without re-estimating the parameters). As model candidates, we considered linear regression models that contained up to 2nd degree terms, and also noise indicator terms (which take values 0 and 1) that effectively adjusts the weights of the non-noisy positions when one or more positions have noisy recordings. Model selection was performed by starting with a base model with one term for each murmur grade prediction, and then terms with low p-values were stepwise added or eliminated (if p>0.05) in order to find the sub model with the lowest bayesian information criterion (BIC) value; a measure of goodness of fit that penalizes high model complexity. After the best model had been estimated by this proceedure, we added, in a similar manner, the noise indicator terms.

To test if using all four recordings improved prediction of AVPGmean, we computed the AUC for prediction of AVPGmean>*u* across a range of thresholds (*u*=7 mm Hg to *u*=30 mm Hg) using the multiple-position model and each of the single-position models. For comparison against the i’th single-position model (the model that utilizes audio from position i to make predictions) we excluded observations with noise in the i’th position.

### Performance Estimation and Model Comparison

Due to the small number of clinically relevant cases of VHD in the dataset, we opted to rely primarily on 8-fold cross validation (CV) to estimate model performance. Extensive use of CV for model selection can result in overestimation of model performance. Prior to performing any data analysis, we therefore set aside a holdout set containing data from 212 (∼10% of the data) participants which contains a total of 91 murmurs of grade 1 or higher, and 55 murmurs grade 2 or higher. All decisions relating to the development of the murmur detection model were done to improve prediction of murmur grade, and not prediction of VHD. Most decisions concerning model architecture and data processing were based on a previous study^20^, thus minimizing the number of modeling decisions made using results from the development set. After the models were developed and results had been obtained, we retrained the murmur detection algorithm using the whole development set, and tested for overfitting on the holdout set.

We hypothesized that neural networks trained to detect the presence of a disease (presence or absence of disease) would outperform the murmur detection algorithm in terms of predicting that disease. AR and MR were the diseases for which there were most training examples (in terms of grade>0), although the disease-threshold had to be set low, to grade 1 or 2, to obtain a substantial number of training examples, thus the positive class contains a large number of low severity cases. For each disease, a network was trained using presence of disease (AR or MR, respectively) as training label, with input data being audio-recordings from the mitral position - the position where the AR or MR associated murmurs are expected to be audible. The algorithms trained directly for disease-prediction used the same model-architecture as the murmur detection algorithm (except for the addition of a classification layer).

### Statistical Analysis

Algorithm performance is measured primarily using AUC, sensitivity, and specificity. AUC is the preferred comparison metric in this study, as it provides a summary of performance that takes into account the rate of both true positives and true negatives, and does not require selection or estimation of a decision threshold. To summarize the overall performance of the murmur detection algorithm across the four auscultation positions, we combine the annotations from all four positions into a single set, and present the AUC for murmur detection performance on this joint set of annotations. All confidence intervals (CI) and p-values reported in this paper correspond to a significance level of 5%, and all statistical tests are two-sided. The level of statistical significance is signified by (*), (**), and (***), which corresponds to p-values within the intervals (0.01,0.05], (0.001,0.01] and (0,0.001] respectively.

We assume that CV-predictions (formed by combining the validation-set predictions for each of the 8 CV-partitions into a single binary vector) are independent and identically distributed, and compute CI’s for sensitivities and specificities using exact methods based on the corresponding binomial distributions. CI’s for AUC are computed under the assumption that the CV-estimates are normally and independently distributed. To prevent biased selection of decision thresholds, these were estimated automatically for each CV-partition set by identifying the threshold that maximized sensitivity+specificity on the CV training-set, subject to the condition that sensitivity should be at least 50%. Selection of optimal decision threshold was performed separately for each prediction target. As measures of interrater agreement, we used linear correlation, and Cohen’s kappa. For Cohen’s kappa we reduced the annotations to binary vectors with 1 indicating annotated murmur grade≥ 2. We also used the murmur-grading of SA to predict which recordings AD had classified as murmur grade≥ 2, and computed the AUC for this prediction.

## RESULTS

### Murmur Prevalence and Annotator Agreement

The prevalence of participants who had a murmur grade≥ 1 in at least one position was 21.4% according to SAs annotations, and 20.1% according to ADs annotations. Increasing the threshold for presence of murmur to grade 2, the prevalences decreased to 13.1% and 13.8% respectively. Murmurs were most prevalent in the aortic position, with 18.6% (CI:16.9%-20.2%) of recordings being graded as 1 or higher (by SA) in the aortic position compared to 11.0% (CI:9.70%-12.4%) in the pulmonic position (**Figure S3** provides a more detailed overview of murmur annotation and prevalence for both annotators). When using SAs annotations to predict which recordings AD had classified as grade≥ 2, an AUC of 0.933 was obtained. The correlation between each set of murmur annotations was 0.83, and Cohen’s kappa was 0.74. When defining presence of murmur as (annotated) grade 2 or higher, we found that 45.8% of the murmurs detected were innocent, in the sense that they did not correspond to at least mild stenosis, or at least moderate regurgitation. The proportion of innocent murmurs decreased to 26.9% when the threshold for regurgitation was lowered to include mild cases. See **Table S2** for a summary statistics concerning the relationship between VHD-grade and presence of murmur.

### Murmur Detection Algorithm Performance

Using the murmur detection algorithm to screen for murmurs of grade≥ 2 on the combined set of recordings from all four positions (effectively treating them as independent samples), an AUC of 0.969 (CI:0.958-0.982), sensitivity of 86.5% (CI:83.2%-89.7%) and specificity of 95.2% (CI:94.7%-95.7%) was obtained. When lowering the murmur grade threshold to include grade 1 murmurs in the murmur category, the AUC decreased to 0.936 (CI:0.931-0.949), see **Figure 2** for ROC curves for cutoff values 1 and 2 respectively. Performance metrics for prediction of murmur grade≥ 2 for each auscultation position can be seen in **Table 1**, which shows that the ability to predict murmurs did not vary notably between positions. The holdout-set AUC for prediction of murmur grade≥ 2 was 0.982 (n=55), and 0.935 for prediction of murmur grade≥ 1(n= 91). As we observed no drop in performance on the hold-out set, we concluded that there was no indication that the murmur detection algorithm was overfitted to the development set.

**Table 1.**
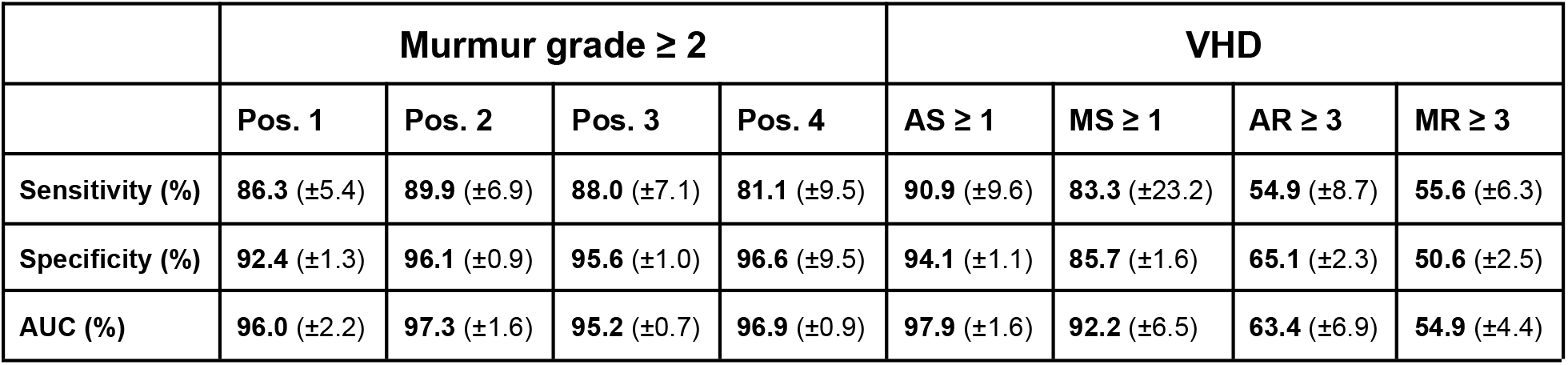
Performance overview of murmur detection algorithm. Performance metrics of the murmur detection algorithm for murmur prediction in different auscultation positions, and for VHD prediction. Values in the AS column were obtained using the multi-position model (described in the Methods chapter), while the other VHDs were predicted using predicted maximum murmur grade across the auscultation positions. 95% confidence intervals are indicated in the parenthesis. VHD = valvular heart disease. AUC = area under the curve.

| | Murmur grade $\geq 2$ | | | | VHD | | | |
| --- | --- | --- | --- | --- | --- | --- | --- | --- |
| | Pos. 1 | Pos. 2 | Pos. 3 | Pos. 4 | AS $\geq 1$ | MS $\geq 1$ | AR $\geq 3$ | MR $\geq 3$ |
| **Sensitivity (%)** | **86.3** ($\pm 5.4$) | **89.9** ($\pm 6.9$) | **88.0** ($\pm 7.1$) | **81.1** ($\pm 9.5$) | **90.9** ($\pm 9.6$) | **83.3** ($\pm 23.2$) | **54.9** ($\pm 8.7$) | **55.6** ($\pm 6.3$) |
| **Specificity (%)** | **92.4** ($\pm 1.3$) | **96.1** ($\pm 0.9$) | **95.6** ($\pm 1.0$) | **96.6** ($\pm 9.5$) | **94.1** ($\pm 1.1$) | **85.7** ($\pm 1.6$) | **65.1** ($\pm 2.3$) | **50.6** ($\pm 2.5$) |
| **AUC (%)** | **96.0** ($\pm 2.2$) | **97.3** ($\pm 1.6$) | **95.2** ($\pm 0.7$) | **96.9** ($\pm 0.9$) | **97.9** ($\pm 1.6$) | **92.2** ($\pm 6.5$) | **63.4** ($\pm 6.9$) | **54.9** ($\pm 4.4$) |

**Figure 2.**
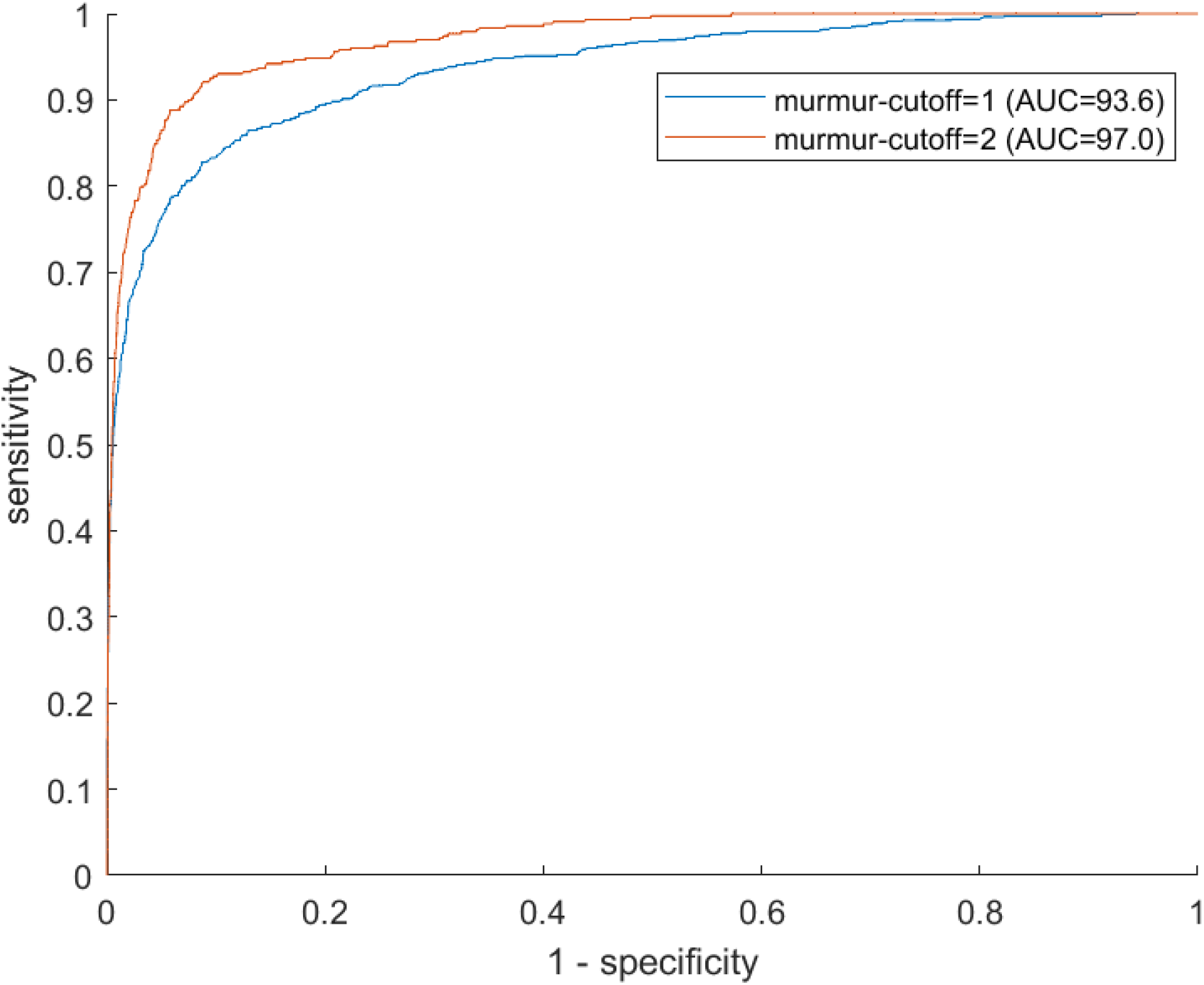
Murmur detection ROC curves. Receiver operator characteristics curves illustrating the algorithm’s ability to predict murmur grade at least 1 (blue) and 2 (red) respectively. Recordings from all four auscultation positions were used in producing the figure.

### VHD Prediction Using Murmur Detection Algorithm

A summary for the murmur detection algorithms ability to predict clinically relevant VHD is presented in **Table 2**, which shows the AUC for prediction of each disease for different disease cutoff values. **Figure S4** shows ROC curves for algorithm prediction of each VHD. Moderate to severe AR was not reliably detected by the algorithm, regardless of which position’s recording was used, with AUC values ranging from 0.576 (mitral position) to 0.634 (using predicted max murmur grade as predictor). Symptomatic AR however, defined herein as presence of angina or dyspnea while walking on flat ground or while resting (currently or previously), was significantly more prevalent in the detected cases than in the missed cases; 16.7% and 1.7% of cases respectively (p=0.02). When the positive class was defined to only include symptomatic AR, sensitivity increased from 54.9% (±8.7%) to 83.3% (±21.1%; 12 cases in total, all of which were severe).

**Table 2.** VHD prediction performance for different positions and disease cutoffs. AUC of murmur detection algorithm for prediction of different diseases, using different disease thresholds *u*, and recordings from different auscultation positions. The bold values indicate the positions associated with the highest AUC (disregarding values in the last row). **\*** The last row shows values obtained using predicted maximum murmur grade (across positions) as VHD-predictor, except for AS which was predicted using the multi-position model. VHD = valvular heart disease. AR = Aortic Regurgitation. MR = Mitral Regurgitation. AS = Aortic Stenosis. MS = Mitral stenosis.

|  | AR |  | MR |  | AS |  | MS |
| --- | --- | --- | --- | --- | --- | --- | --- |
| | $u=3$ | $u=4$ | $u=3$ | $u=4$ | $u=1$ | $u=2$ | $u=1$ |
| Pos. 1 | **62.2** ($\pm 6.8$) | **68.1** ($\pm 5.1$) | 52.5 ($\pm 4.1$) | 57.3 ($\pm 8.1$) | 96.7 ($\pm 1.9$) | 98.4 ($\pm 2.1$) | 89.0 ($\pm 10.9$) |
| Pos. 2 | 59.6 ($\pm 8.0$) | 62.1 ($\pm 11.3$) | **56.3** ($\pm 3.1$) | 57.6 ($\pm 13.5$) | **98.2** ($\pm 1.0$) | **98.7** ($\pm 0.5$) | **89.6** ($\pm 8.8$) |
| Pos. 3 | 58.4 ($\pm 6.8$) | 58.7 ($\pm 7.6$) | 54.9 ($\pm 3.3$) | **61.3** ($\pm 10.4$) | 97.0 ($\pm 1.5$) | 98.4 ($\pm 0.8$) | 85.4 ($\pm 17.6$) |
| pos. 4 | 57.6 ($\pm 6.0$) | 56.1 ($\pm 10.7$) | 55.8 ($\pm 3.9$) | 55.8 ($\pm 11.0$) | 92.1 ($\pm 4.5$) | 97.6 ($\pm 1.5$) | 86.0 ($\pm 15.5$) |
| Prediction based on all positions * | 63.4 ($\pm 6.9$) | 67.3 ($\pm 8.7$) | 54.9 ($\pm 4.4$) | 57.8 ($\pm 6.9$) | 97.9 ($\pm 1.4$) | 99.3 ($\pm 0.4$) | 92.7 ($\pm 7.8$) |

MR was the least reliably detected VHD, as moderate to severe MR was predicted with an AUC of only 54.9% (CI:50.6%-59.3%) using predicted maximum murmur grade as predictor, or 55.8% when basing prediction on the mitral recording. No consistent trends with regards to which position was most predictive of MR was observed. As was the case for AR, detectability of MR was highly influenced by the presence of symptoms. Symptomatic MR, defined the same way as it was for AR, was significantly more prevalent in the detected cases than in the missed cases; 12.9% and 4.5% of cases respectively (p=0.02). The algorithm detected 22 out of 27 (81.5% ±15.9%) symptomatic cases of moderate to severe MR, which is a considerable increase in detection rate relative to 55.6% (±6.3%) when asymptomatic cases were included.

Presence of MS was predicted with sensitivity 83.3% (CI:51.6%-98.0%), specificity 85.7% (CI:84.0%-87.2%), and AUC 0.922 (CI:0.858-0.987). None of the murmurs observed for the MS-cases were diastolic, despite the characteristic MS-murmur occurring during diastole. 7 of the total 13 cases (53.9%) of MS also had moderate to severe AS, indicating a considerable overlap between the two diseases. Of the 10 detected MS-cases, 4 had no presence of AS, and 3 were not associated with any other VHD (excluding trace and mild levels of AR and MR, which were common in the dataset).

We hypothesized that training on the disease labels directly might allow the networks to detect more subtle disease features, and thereby improve performance. Training the networks to differentiate moderate to severe AR directly failed to improve prediction significantly beyond what was achieved with the murmur detection algorithm, as it predicted moderate to severe AR with an AUC of only 55.9% (CI:45.4-66.5), slightly, but not significantly, lower than that achieved using the murmur detection algorithm. Similarly, the network trained to predict moderate to severe MR achieved an AUC of only 57.5% (CI:52.3-62.6), which is slightly higher, but not significantly different from the murmur detection algorithms AUC of 55.8%.

Mild or greater AS was predicted by the multi-position model with AUC 0.979 (CI:0.963-0.995), sensitivity 90.9% (CI:78.3-97.5%) and specificity 94.5% (CI:93.3-95.5%), whereas moderate to severe AS was predicted with AUC 0.993 (CI:0.972-0.996). In comparison, maximum (predicted) murmur grade predicted mild or greater AS with AUC 0.972, and moderate or greater AS with AUC 0.991. Accuracy was in general higher for more severe AS, as can be seen in **Table 2**, or in **Figure 6** which shows AUC as a function of AVPGmean cutoff threshold. A positive test using the multi-position model increased the risk of mild or greater AS from 2.12% to 19.1% (assuming an unselected population of individuals aged 40 or higher). For moderate to severe AS, the corresponding values were 1.27% and 17.1%. A negative result was estimated to rule out presence of AS with >99% probability.

The pulmonic position was the position that produced the most accurate prediction of both at least mild and at least moderate AS (see **Table 2** which shows prediction performance associated with each position), with an associated AUC value of 0.982 (±0.010) for detection of mild or greater AS. In contrast, prediction based on the aortic position achieved a lower AUC of 0.967, although the difference was not significant (p=0.16). Screening for mild or greater AS on the holdout set using the multi-position model, all 6 cases were detected, the specificity was 90.6%, and the AUC was 0.988.

Using the multi-position model to predict at least mild AS resulted in 4 missed cases (of 45 cases), of which 3 were mild, and one was moderate (AVPGmean = 27.17 mm Hg). There were 102 false positive predictions, but 20.1% of these observations had AVPGmean>10 mm Hg, which may be of clinical interest despite falling below the set threshold.

### Joint Screening for Clinically Relevant Cases

As there is overlap between the different types of VHD, a false positive test for a specific VHD may nevertheless result in appropriate clinical follow up. To evaluate the overall screening potential of the algorithm, we screened jointly for clinically significant cases using predicted maximum murmur grade as predictor, and defined the positive class as the set of participants with at least moderate regurgitation or at least mild stenosis. These cases were detected with sensitivity 54.1%, specificity 60.5%, and AUC 0.61 (CI:0.567-0.653). In the process, 97.7% (of 44 cases) of AS cases, all 12 MS cases, 58.7% of AR cases and 48.8% of MR cases were detected. When we included only severe cases of regurgitation in the positive class, the algorithm detected 67.2% of AR cases, 51.8% of MR cases, and achieved an overall AUC of 0.731 (CI:0.669-0.793). When we instead included only symptomatic cases (dyspnea or angina) of AR and MR from the positive class, the AUC increased considerably to 0.912 (CI:0.891-0.933; 44 of 60 detected). **Figure 3** shows ROC curves for prediction of these different clinical categories, and **Figure 4** the proportions of different clinical subgroups that were identified as well as the prevalence of false positive predictions. When screening for at least moderate regurgitation or mild or greater stenosis on the holdout set, the algorithm detected all 6 cases of presence of AS, 1 of 1 cases of MS, 9 of 15 cases of AR, 15 of 36 cases MR, and achieved an AUC 0.652 for overall detection of significant cases.

**Figure 3.**
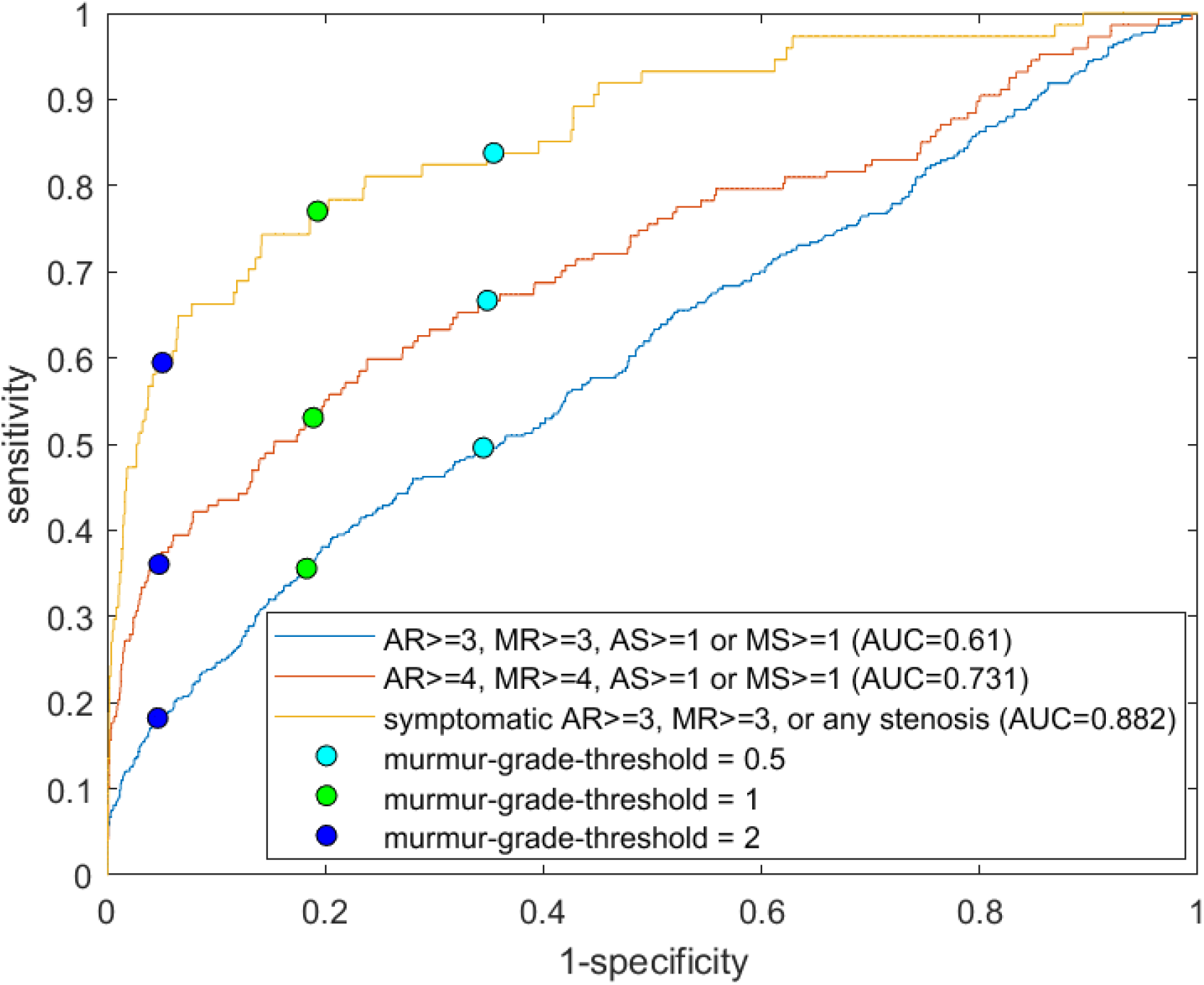
ROC curves for prediction of clinically relevant cases. Each receiver operating characteristics curve corresponds to prediction of a different definition of clinically relevant VHD, with definitions shown in the Figure legend. For the yellow curve, asymptomatic cases of regurgitation were excluded from the positive class. The circles show sensitivities and specificities for murmur grade decision thresholds of 0.5, 1.0 and 2.0 respectively. The position with the highest predicted murmur grade was used as the predictor. VHD = valvular heart disease. AR = Aortic Regurgitation. MR = Mitral Regurgitation. AS = Aortic Stenosis. MS = Mitral stenosis.

**Figure 4.**
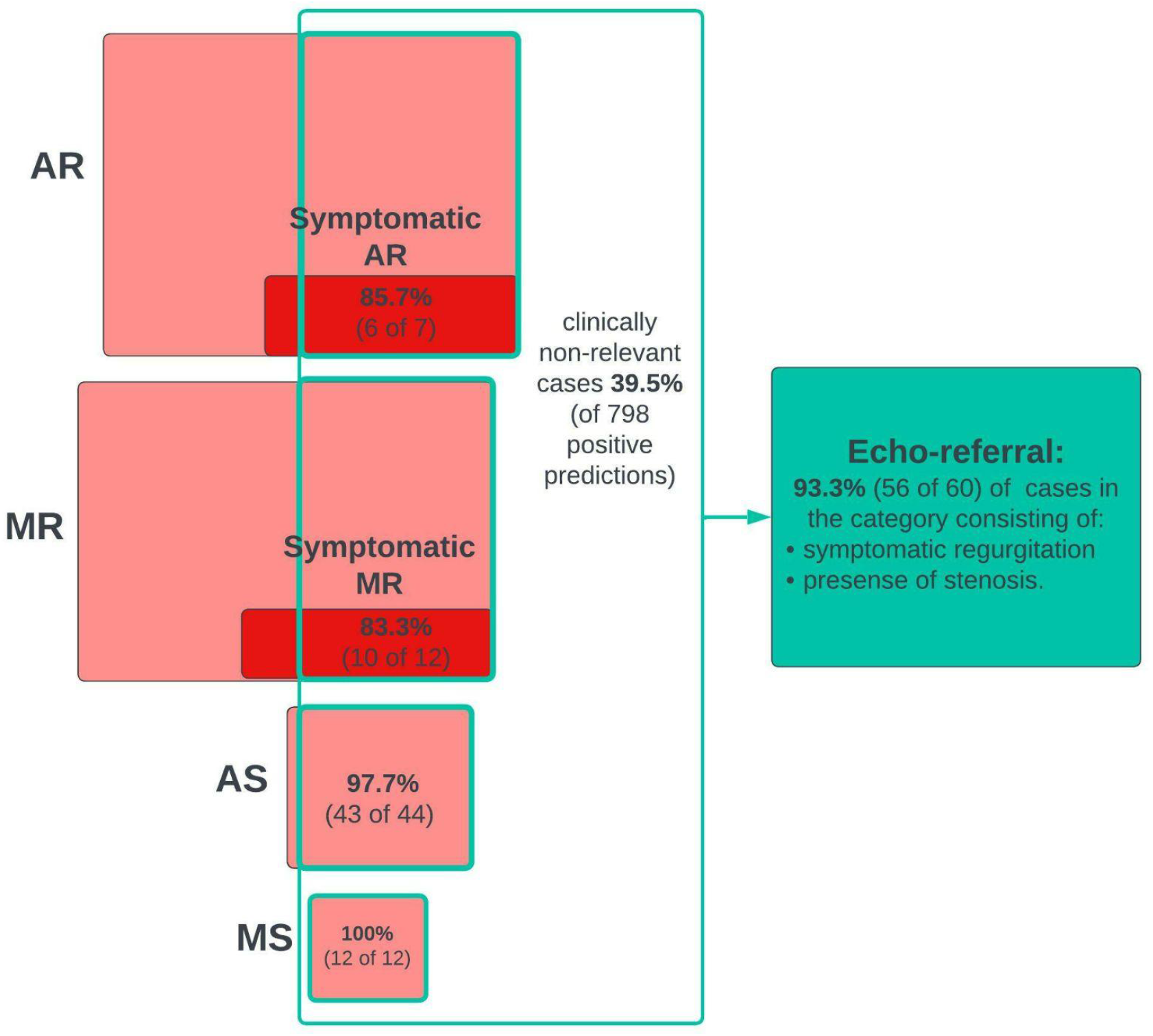
Joint screening for clinically relevant cases. Diagram that shows numbers and proportions of clinically significant cases detected in the process of screening jointly for all clinical cases using the murmur detection algorithm, as well as the prevalence of false positive predictions. Predicted maximum murmur grade was used as the predictor of clinical cases. A positive prediction was counted as a true positive if the participant had a significant grade for at least one VHD. Included in the pathological VHD categories are mild or greater stenosis and at least moderate regurgitation. The red boxes represent subgroups of participants who had or used to have symptoms (angina, or dyspnea while walking calmly on a flat surface or resting). VHD = valvular heart disease. AR = Aortic Regurgitation. MR = Mitral Regurgitation. AS = Aortic Stenosis. MS = Mitral stenosis.

### Multi-position Prediction of Aortic Valve Mean Pressure Gradient

The multi-position linear regression model for prediction of AVPGmean obtained after model selection and parameter estimation was

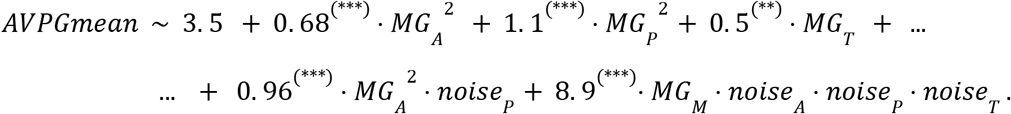

The subindeces *A,P,T,M*refers to the aortic, pulmonic, tricuspid and mitral position respectively, *MG*_*i*_ denotes the predicted murmur grade of position *i*, and *noise*_*i*_is the indicator variable that modifies the murmur grade weights if the recording from position *i* is noisy (in which case *MG*_*i*_ is set to zero). All parameters p-values were lower than 0.01.

After selecting the model, we performed cross-validation to estimate how well the multi-position model predicted AVPGmean compared to the single-position models across a range of cutoff thresholds, using AUC as comparison metrics. **Figure 5** shows the results for each position, with significant differences indicated by **\***. For AVPGmean cut off 10 mm Hg, the multi-position prediction outperforms each of the single-position predictions by amounts corresponding to p-values 0.09, 0.02, 0.40 and 0.01 respectively.

**Figure 5.**
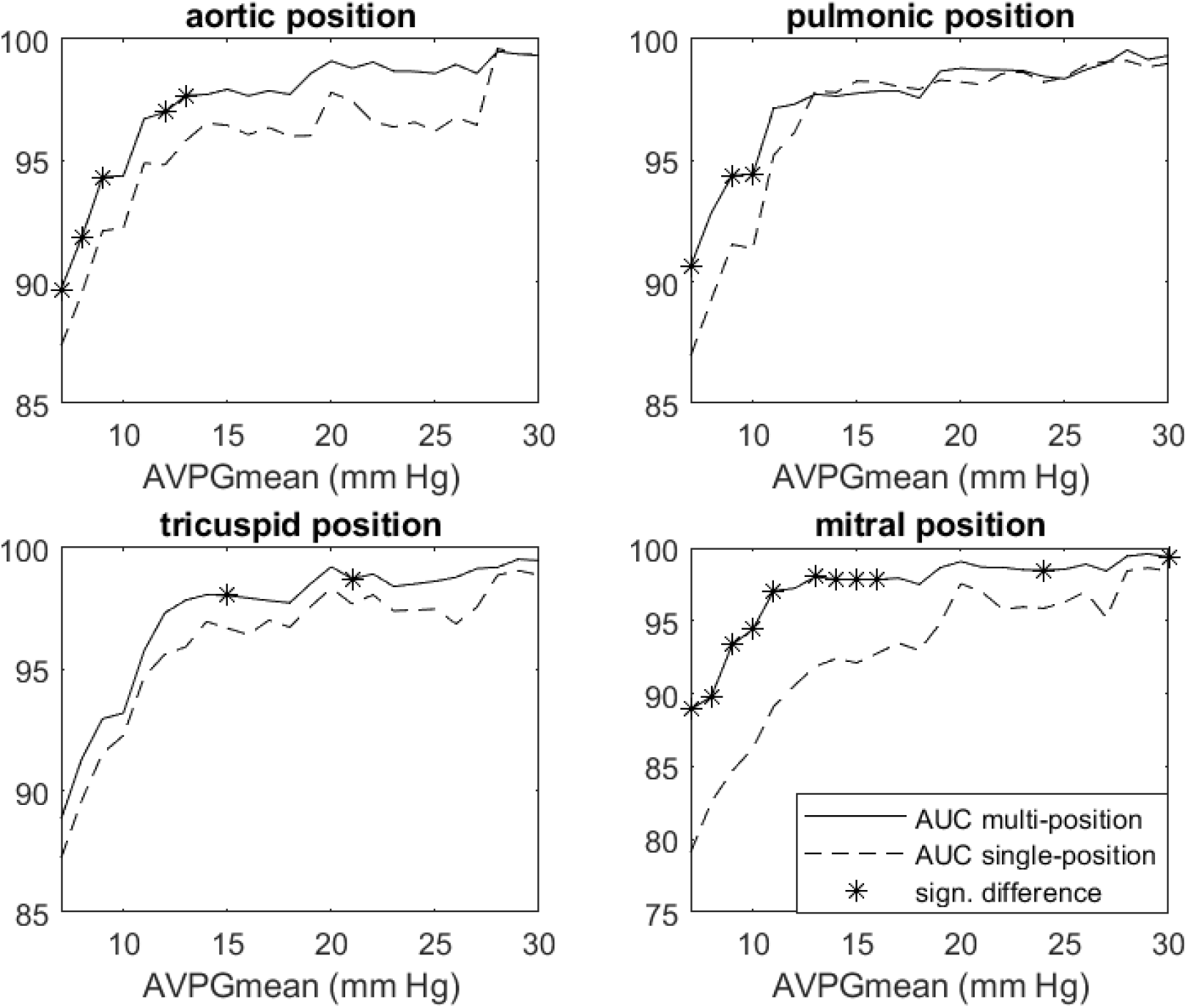
Multi-position model vs single-position AUC for AVPGmean prediction. Each panel shows AUC for prediction of cases where AVPGmean > *u* for thresholds *u* ranging from 7 mm Hg to 30 mm Hg for the single-position (dashed line) and multi-position (solid line) model respectively. Each single-position model uses only data from the index position to make predictions, whereas the multi-position model uses data from all four positions. Significant differences are marked with **\***. AUC = area under the curve. AVPGmean = aortic valve mean pressure gradient.

**Figure 6.**
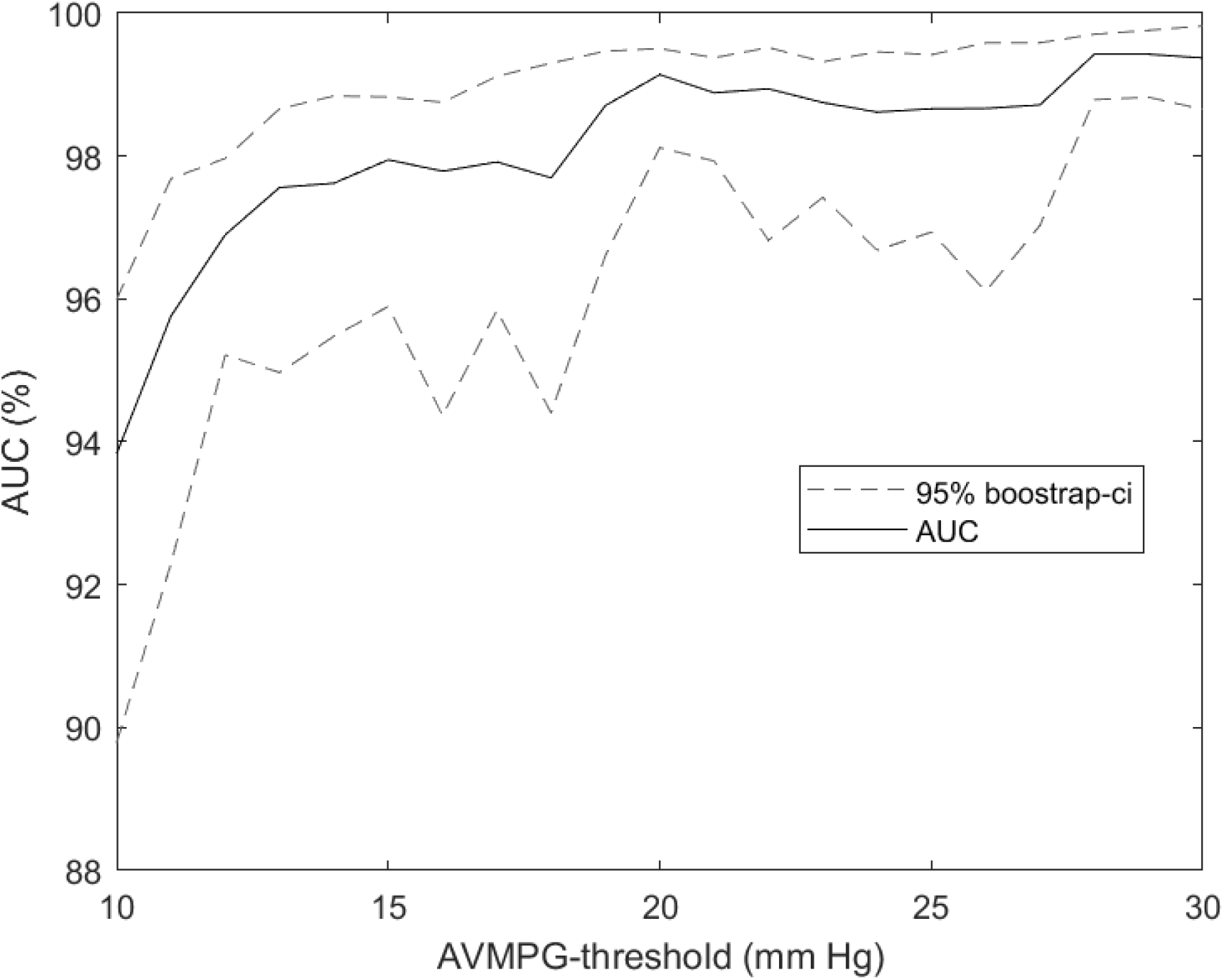
AUC as a function of aortic stenosis cutoff threshold. AUC for prediction of AVPGmean>*u* across a range of thresholds *u*, using the multi-position model on the subset with noise in up to 3 out of 4 auscultation positions. Confidence intervals were obtained using bootstrapping. Recordings annotated as noisy in all four positions were excluded from the analysis. AVPGmean = aortic valve mean pressure gradient. AUC = area under the curve.

### VHD Prediction Using Multiple Predictors

An overview of the multivariate logistic regression models can be seen in **Table 3**, which also shows the AUCs for prediction of each VHD (as defined by severity grade cutoff values). The estimated coefficients of each model can be seen in **Table 4**.

**Table 3.** Multivariate regression models that include clinical variables. The table shows the variables that were included in the logistic regression models that use clinical factors and predicted murmur grades to predict VHD, as well as the AUC for each model. Interaction between variables is denoted by “: ”. As auscultation based prediction input, the AR and MS models use maximum murmur grade, the MR model uses the mitral position murmur grade, and the AS model uses the output from the multi-position model. VHD = valvular heart disease. AR = Aortic Regurgitation. MR = Mitral Regurgitation. AS = Aortic Stenosis. MS = Mitral stenosis.

| Prediction target | AUC | Model |
| --- | --- | --- |
| AR≥3 | **0.747**<br>($\pm 0.035$) | AR ~ murmur grade + age + sex + dyspnea uphill + heart rate |
| MR≥3 | **0.672**<br>($\pm 0.039$) | MR ~ murmur grade + age + heart rate |
| AS≥1 | **0.982**<br>($\pm 0.019$) | AS ~ murmur grade + sex + sex : murmur grade |
| MS≥1 | **0.938**<br>($\pm 0.041$) | MS ~ murmur grade + age + heart rate |

**Table 4.** Estimated parameters of clinical variable models. The table shows the estimated coefficients of the variables included in the multivariable logistic regression models. The *murmur grade* column shows the effect of the explanatory variable derived from predicted the murmur grade(s); the AR and MS models use predicted maximum murmur grade, the MR model uses the mitral position predicted murmur grade, and the AS model uses the output from the multi-position model. Statistical significance of the estimated parameters is marked by (*), (**), and (***), which indicates p-values within the intervals (0.01-0.05], (0.001-0.01] and (0-0.001] respectively. VHD = valvular heart disease. AR = Aortic Regurgitation. MR = Mitral Regurgitation. AS = Aortic Stenosis. MS = Mitral stenosis.

| prediction target | age | murmur grade | murmur grade : sex | sex | dyspnea | heart rate |
| --- | --- | --- | --- | --- | --- | --- |
| AR≥3 | 0.0034*** | 0.0390*** | - | 0.0330* | 0.0270* | -0.0017** |
| MR≥3 | 0.0052*** | 0.0120 | - | - | - | -0.0033*** |
| AS≥1 | - | 0.0247*** | 0.0248*** | -0.0848*** | | |
| MS≥1 | 0.0004* | 0.0200*** | - | - | - | 0.0008*** |

Prediction performance increased significantly for AR (p=0.002) and MR (p=0.007) compared to prediction based only on murmur grade alone when including additional clinical variables into a multivariate logistic regression model. Prediction of both mild or greater AS and MS was improved slightly, but not significantly, with AUCs increasing from 0.979 to 0.982 (p=0.30), and from 0.922 to 0.938 (p=0.33) respectively. At least moderate AS was detected with AUC 0.995 (CI:0.991-0.998, up by 0.002, p=0.17), sensitivity 96.3% (CI:81.0%-96.3%) and specificity 96.7% (CI:95.7%-97.4%). At least mild AS was detected with sensitivity 88.6%, and specificity 95.0%, and of the false positives, 35.9% had either at least moderate AR, at least moderate MR, or AVPGmean>10 mm HG.

In all models, age and heart rate were significant predictors, except in the AS model, where they accordingly were not included as model parameters. Interaction between sex and murmur grade was highly significant (p-value<0.0001), indicating that for men, a high murmur grade is a particularly big risk factor for AS.

## DISCUSSION

The murmur detection algorithm was highly accurate at predicting AS. We observed a clear trend of higher detectability for more severe forms of the disease, but even mild cases were detected well. The multi-position model was the overall preferable model for predicting AS. The pulmonic position was comparable in terms of predicting AVPGmean in the higher range (AVPGmean>15 mm Hg) when restricting the comparison to the subset with clean pulmonic recordings. However, the multi-position model was significantly more accurate at predicting AVPGmean<10 mm Hg, and in general appeared to predict the pressure gradient more accurately in the lower range. Furthermore, it is not limited to cases with clean pulmonic or aortic recordings, and extending prediction of AS to the set of observations with noise in up to 3 positions did not result in any notable loss in accuracy. A theoretical motivation for preferring the multi-position model is that taking a weighted average may help dampen the random variability in the predictions of each position. Indeed, a trend we observed in our study was that the average of annotated murmur grades was more predictive of AS than individual annotations. When we computed the AUC for prediction of AS based either on individually annotated murmur grade or on the average annotated murmur grade, we found that the AUC of the annotator average improved upon the AUC of the individual annotations (using the highest AUC of the two annotators for comparison) by 1.7%, 0.1%, 2.8%, and 2.9% for positions 1-4 respectively.

The accuracy with which our algorithm detected AS compares favorably against metrics on clinician accuracy we found in the litterature. In a study by Jaffe et al.^33^, severe AS was detected only with a sensitivity of 83% and specificity of 79%. In a 2021 study by Chorba et. al.^34^ 3 expert cardiologists annotated recordings primarily from the aortic position with presence or absence of murmur, which were subsequently used to predict presence of moderate to severe AS in a selected cohort consisting of confirmed pathological cases and healthy controls. The highest performance (in terms of high SN+SP) achieved via this prediction scheme was a sensitivity of 82.5% (CI:69.6%-93.6%) and specificity of 90.2% (CI:83.1%-96.3%), and the average sensitivity and specificity across annotators was 90.0% and 71.1% respectively.

The aforementioned study by Chorba et al. offers a particularly interesting comparison to our study. They trained a deep convolutional neural network to predict murmur grade, achieving an AUC of 0.958 for predicting murmurs of grade 2 or higher, which we outperform with an AUC of 0.969 (±0.12), although the difference could be influenced by differences related to annotators or test method (they used a test set, whereas we used cross-validation for model testing).

Furthermore, they achieved sensitivity 93.2% (CI:86.9%–98.5%), specificity 86.0% (CI: 80.9%–91.0%), and AUC 0.952 for prediction of moderate to severe AS based preferentially on the aortic recording, which is similar to our AUC of 0.967 when using the aortic recording for prediction. We improved prediction of AS further by combining all four predicted murmurs in the multi-position model, obtaining AUC’s of 0.978 (CI:0.961-0.994) and 0.992 (CI:0.988-0.997) for prediction of at least mild and at least moderate AS respectively, thus significantly outperforming their results. We note that Chorba et al. preferred for prediction of AS to use the aortic recording (which is often recommended for detecting the AS-murmur), using the pulmonic position as a secondary choice, yet our results suggests that the pulmonic position recording might be more suitable for prediction of at least mild (p=0.16) or at least moderate (p=0.82) AS.

A caveat to the above comparison is that Chorba et al. did not specify the exact criteria they used for grading AS beyond stating that they followed the American Society of Echocardiography’s Guidelines, thus differences in performance could be impacted by differences in how AS was graded. Defining AS in terms of the AVPGmean could result in a target that is easier to predict, as it excludes cases of AS where the AVPGmean is low due to low cardiac output, and such cases might be harder to detect. However, we believe such cases are rare and unlikely to impact classification substantially^13^, and since the performance increase was significant even when using a low disease-threshold, we believe it reflects genuine methodological improvement.

While AS was detected with comparable accuracy in the present and the Chorba et al.’s study, results for detection of at least moderate MR were dramatically different, as they predicted these cases with an AUC of 0.865 (29 cases, 62 healthy controlls), which exceeds our result (mitral position prediction) of 0.558 (±0.039) with a huge margin. The discrepancy is unlikely to be due to differences in model performance, so this wide performance gap suggests that cases of moderate to severe MR in selected cohorts are indeed more discernible than in unselected cohorts. Consistent with this observation is the fact that detection rates for MR went up significantly within our own study when asymptomatic cases were excluded from the positive class, with a similar but even more notable trend for detection of AR. Another study which supports this conclusion is a 2018 study by Gardezi et. al.^9^, in which they tested the accuracy of auscultation in a cohort of asymptomatic patients aged 65>years, and found that diagnosis of significant VHD by auscultation was not significantly better than chance. Myerson et al. found similar results in their 2017 study of unselected and asymptomatic participants aged 65 or older with no previous VHD diagnosis, where GPs identified significant VHD (moderate to severe regurgitation or at least mild stenosis) with AUC of only 0.56^35^.

### Clinical Implications

The fact that AS was detected reliably in a cohort representative of the general population has promising health care implications. AS, whether it is asymptomatic or not, is associated with poor prognosis when left untreated, and presence of symptoms are not reliable indicators of disease-severity. The need for an accurate and cost-effective screening tool is further emphasized by the high prevalence of undiagnosed AS in the elderly population, especially when considering that effective treatment exists. L. d’Arcy et al. found a 1.3% prevalence of newly diagnosed AS (defined as aortic valve thickening or calcification with a maximum aortic transvalvular velocity>2.5 m/s) in a general cohort consisting of individuals aged≥ 65 years^15^. Extrapolating from these numbers (assuming that ours and their definition of presence of AS matches reasonably well), we estimate that given a population size of 10,000 elderly individuals, our multivariate risk-factor algorithm would detect 115 of the expected 130 individuals with undiagnosed AS, whilst producing ∼495 false positives. If we reclassify 35.9% of false positives as true positives as they are expected to be of clinical interest (see the section on the clinical factor models), the expected number of false positives drops to 178.

Beyond diagnosing AS, there is data that suggests that a murmur detection algorithm could help identifying high risk subgroups not typically considered for auscultation. A recent 35-year follow-up study demonstrated that murmur of grade 2 or higher in seemingly healthy middle aged men with moderate-grade murmurs had an 89.3-fold (CI: 39.2-211.2) age-adjusted risk of undergoing aortic-valve-replacement later in life, and a 1.5-fold (CI:0.8-2.5) age-adjusted increased risk of CVD death^36^. Thus, even in subgroups not typically considered for cardiac auscultation, screening for murmurs could provide clinically valuable information.

### Study and Algorithm Strengths and Limitations

A limitation of the murmur detection algorithm is that it does not automatically screen for noise, which means that the user must decide which recordings are of sufficient quality. This is likely not an issue if the user is a clinician, but could potentially be an issue if the user is a patient performing the procedure in their home. Another limitation is that features other than murmur grade that might be of diagnostic value, such as features relating to timing or pitch, have effectively been ignored. The diagnostic accuracy with which the algorithm detects a disease is therefore limited by the degree to which the disease is associated with murmur grade.

The main limitations of this study are related to the highly imbalanced dataset and subsequent decision to use cross-validation for producing the main results rather than a holdout set. In particular, the performance of the risk-factor models are at risk of being inflated, as they were tuned to predict a very small number of pathological cases. Claims about which positions or methods are optimal for predicting AS need to be tested on an external dataset, as the small number of cases results in low statistical power, leaving many potentially interesting results in need of verification. We were also limited in terms of model development, as we had to restrict ourselves to exploring a small set of simple models in order to limit risk of overfitting. Finally, we note that the recordings used in this study were collected in a setting that was less noisy than can be expected in a rushed acute clinical setting, and further work may be required to improve robustness to such noise, unless quality control can be used.

A strength of the algorithm is that we have demonstrated its accuracy on a dataset containing samples with noise in up to 3 positions, and it utilizes information from all four standard positions in order to produce more robust information for audio-segmentation. Only 1.4% of the dataset was excluded from analysis due to all recordings containing noise, so the performance metrics we present are unlikely to be substantially inflated due to exclusion of noisy samples (except the model that exclusively predicted MR, whiched used the mitral position). Another strength of the algorithm is the relative simplicity of the model used to predict murmur grade, which entails faster computation. We also believe its simplicity puts it at lower risk of overfitting than deeper neural networks that are often employed in automated heart sound analysis research, as their enhanced flexibility and adaptability might increase their risk of adapting to features that are specific to the development set, such as the type of stethoscope used.

The main novelty and strength of our study lies in the unfiltered nature of the dataset, which to the best of our knowledge makes this the first study of its kind to utilize such data for both model training and testing, thus allowing us a unique opportunity to get a more realistic assessment of the ability of automated HS analysis to screen for VHD in general populations. We also had access to murmur annotation, ground truth results from echocardiography, and data on various clinical factors, which to our knowledge is a novel combination of data for this kind of study.

Finally, the HS annotations used in this study were obtained using a rigorous procedure in which considerable effort was put into ensuring consistent data annotation and high interrater agreement, and we believe this has resulted in high quality training data which in turn contributes to high algorithm prediction performance.

## CONCLUSIONS

In this study we developed a murmur detection algorithm which detects mild or greater AS in an unselected cohort with an accuracy similar to or exceeding previously reported metrics from similar studies that were based on selected cohorts. Prediction accuracy of AS benefited from using a model that utilized audio from all four standard auscultation positions, achieving high accuracy on a dataset which excluded only the samples that had noisy audio in all four positions. AR and MR were not reliably detected, but detection accuracy increased significantly when only symptomatic cases were targeted, and when age, sex, heart rate, and dyspnea were included as predictors. Our results indicate that automated HS analysis could be a highly cost effective screening tool for VHD, and could considerably reduce the number of undetected clinically important cases in the general population.

## Supporting information

STARD 2015 checklist

COI disclosure

## Data Availability

The Tromso Study data is not publicly available, but researchers can apply for access at: https://uit.no/research/tromsostudy

## List of non-standard Abbreviations

AVPGmean: aortic valve mean pressure gradient
AUC: area under the receiver operating curve
HS: heart sound
VHD: ventricular heart disease
GP: general practitioner
CV: cross validation
AR: aortic regurgitation
MR: mitral regurgitation
AS: aortic stenosis
MS: mitral stenosis

## ARTICLE INFORMATION

### DATA AND CODE AVAILABILITY

The Tromsø Study data is not publicly available, but researchers can apply for access at: https://uit.no/research/tromsostudy

Descriptions of the questionnaire variables used in this study can be found at: http://tromsoundersokelsen.uit.no/tromso/

The code is open sourced using the The GNU Affero General Public License. It is available at: https://github.com/uit-hdl/heart-sound-classification

## SUPPLEMENTARY MATERIALS

Tables S1-S3

Figures S1-S4

Comparison Between Continuous and Binary Labels

