## Supplementary Materials for "Algorithm for Predicting Valvular Heart Disease from Heart Sounds in an Unselected Cohort"

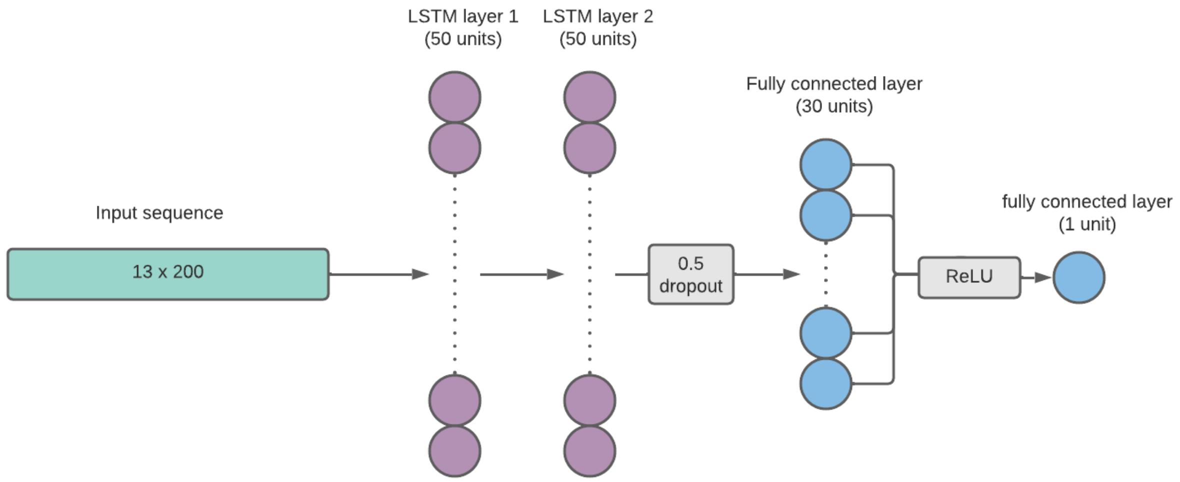


**Figure S1. Network Architecture**

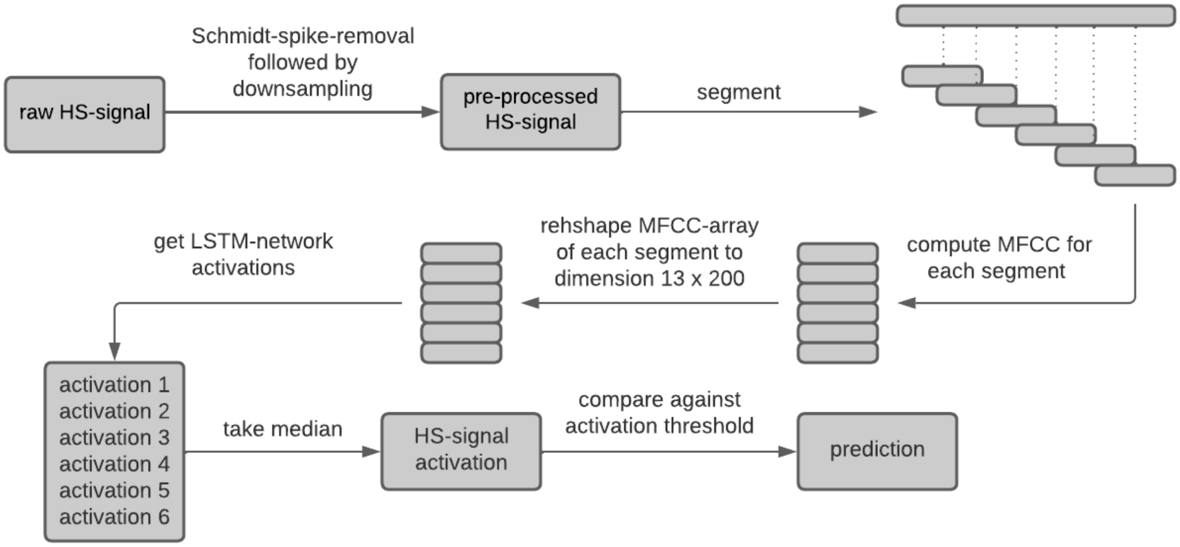


**Figure S2.    Processing steps between raw heart sound input and algorithm prediction.**

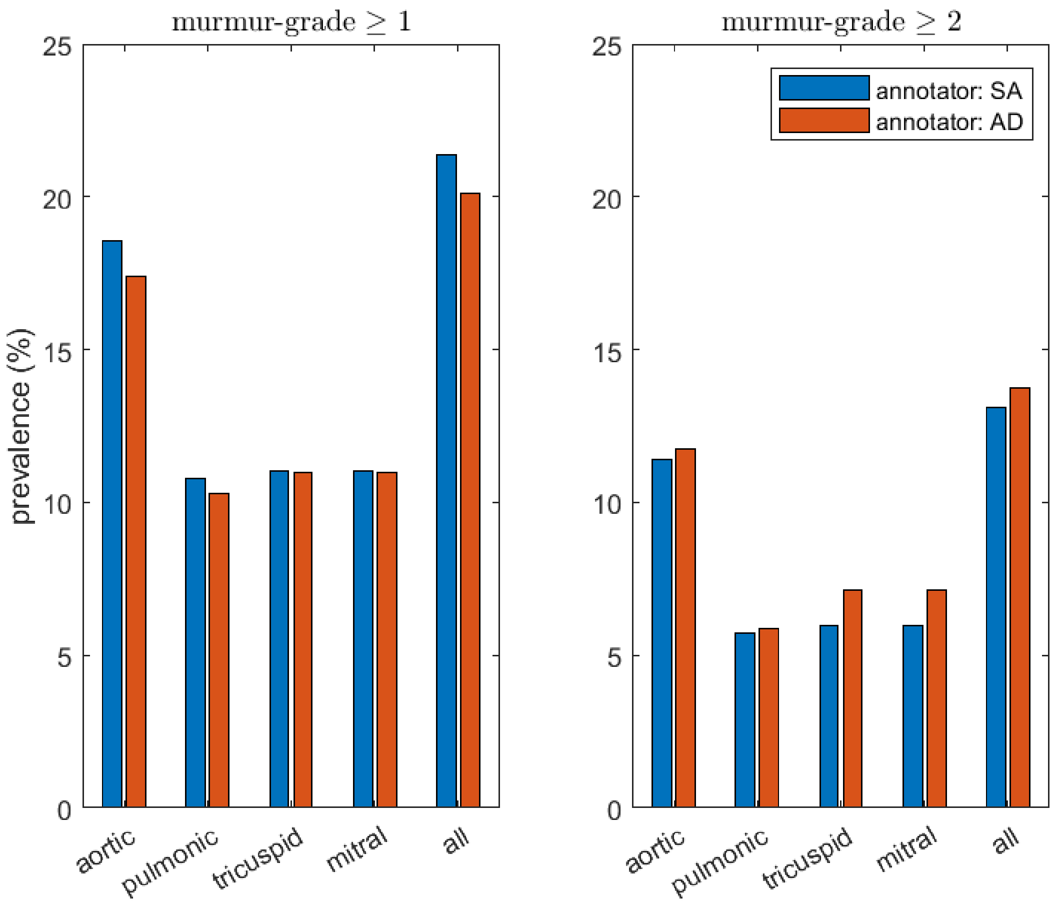


**Figure S3. Prevalence of murmurs as perceived by each annotator.**
The left panel shows the percentage of recordings for which each annotator (annotator names are abbreviated to SA and AD) perceived a murmur of grade at least 1. The panel to the right shows the same information, but with grade 2 as threshold. The bars furthest to the right (labelled “all”) show the prevalence of observations for which at least one auscultation position had a murmur grade that exceeded the cutoff threshold - i.e. it considers the maximum murmur grade across the recordings of a study participant.

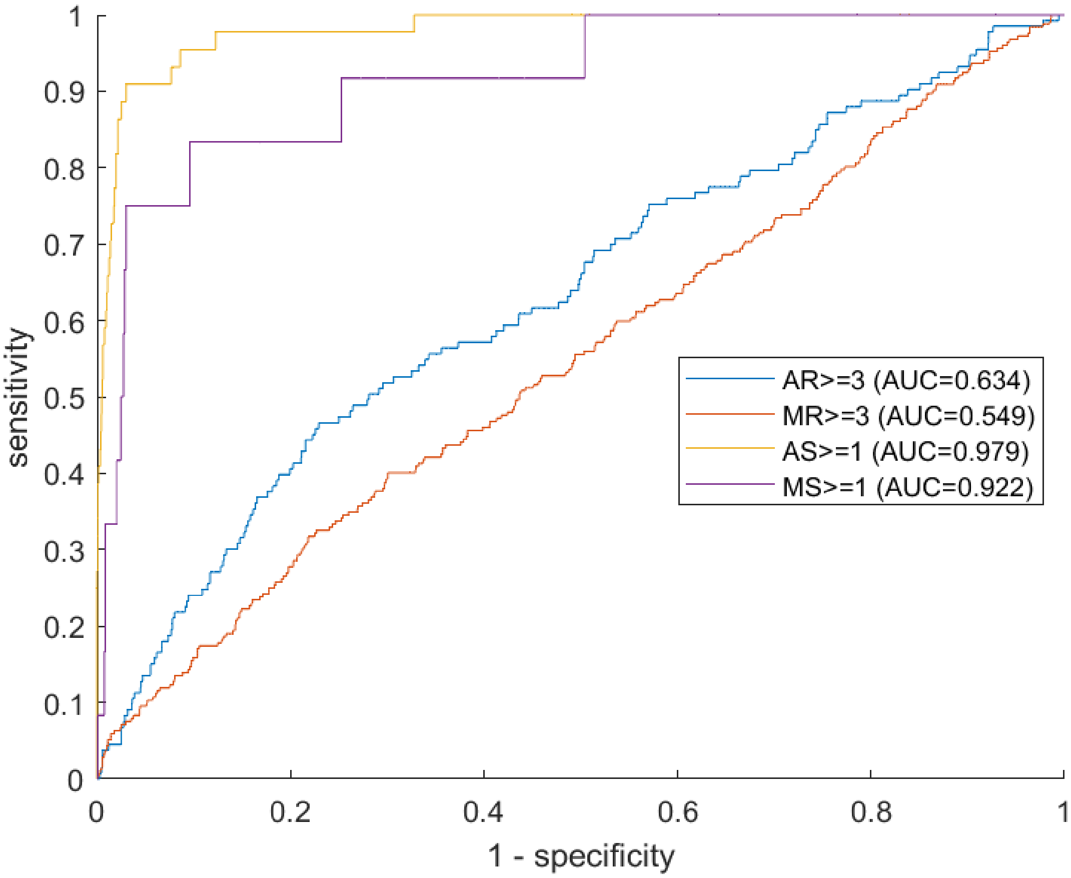


**Figure S4. ROC curves for prediction of each VHD**

The figure provides an overview of the algorithm's ability to predict each VHD, the presence of which is defined using the severity grade relative to a fixed threshold (indicated in the legend).

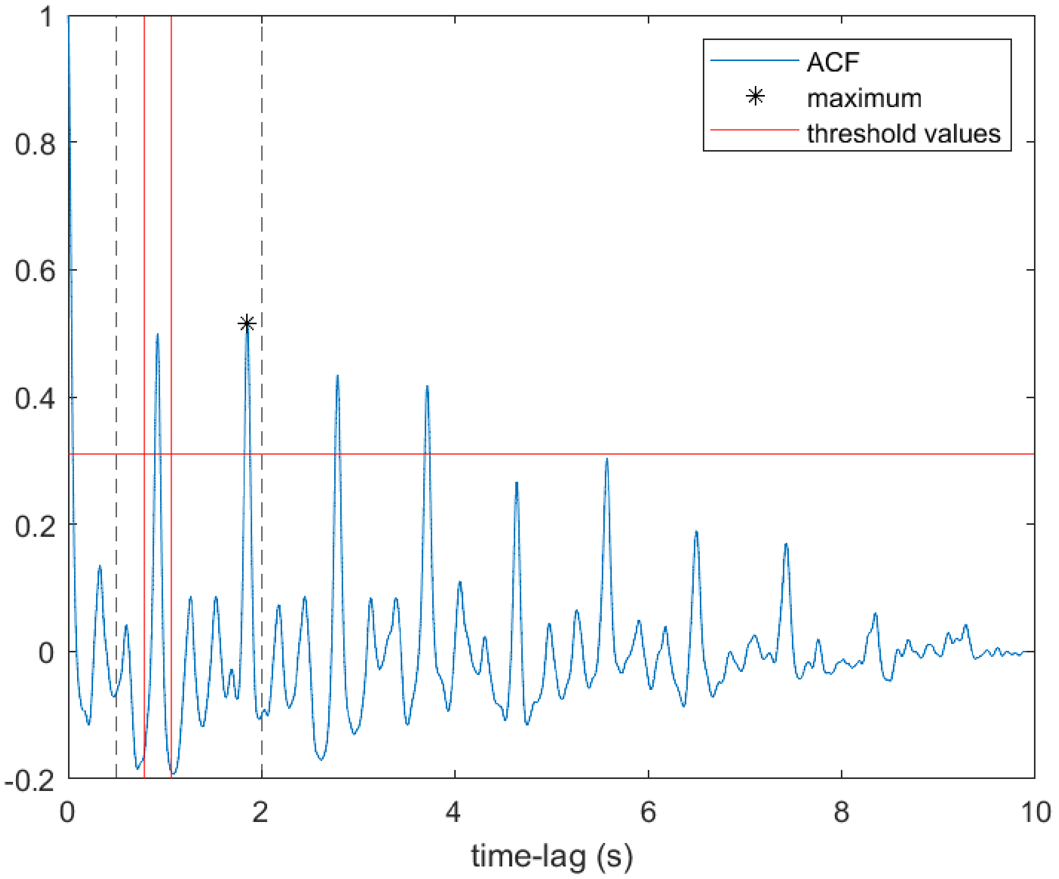


**Figure S5.** **Estimation of heart rate from the auctocorrelation function**
Example of how the modified version of Springer's segmentation algorithm identifies the heart rate peak in the autocorrelation function corresponding to the heart sound audio. Threshold values are calculated based on the largest peak within the search interval [0.5s, 2s].

| VHD prevalence | | | | |
|---|---|---|---|---|
| | AR | MR | AS | MS |
| grade 1 | 205 (9.7%) | 273 (12.9%) | 19 (0.9%) | 10 (0.5%) |
| grade 2 | 106 (5%) | 513 (24.2%) | 26 (1.2%) | 2 (0.09%) |
| grade 3 | 84 (4%) | 223 (10.5%) | 6 (0.28%) | 1 (0.05%) |
| grade 4 | 66 (3.1%) | 69 (3.2%) | - | - |
| **Sex and age distribution** | | | | |
| | AR≥3 | MR≥3 | AS≥1 | MS≥1 |
| female, n | 66 (46.9%) | 145 (51.8%) | 16 (31.4%) | 10 (76.9%) |
| male, n | 84 (53.1%) | 147 (48.2%) | 35 (68.6%) | 3 (23.1%) |
| mean age (SD) | 71.3 (0.73) | 69 (0.6) | 74.9 (1.9) | 77.7 (1.4) |
| 40-49, n | 5 (3.3%) | 17 (5.8%) | 0 (0%) | 0 (0%) |
| 50-59, n | 8 (5.3%) | 33 (11.3%) | 1 (2.0%) | 0 (0%) |
| 60-69, n | 39 (26%) | 83 (28.4%) | 11 (21.6%) | 2 (15.4%) |
| 70-79, n | 73 (48.7%) | 119 (40.8%) | 22 (43.1%) | 5 (38.5%) |
| >80, n | 25 (16.7%) | 40 (13.7%) | 17 (33.3%) | 6 (46.2%) |
| **Questionaire data and clinical variables** | | | | |
| | AR≥3 | MR≥3 | AS≥1 | MS≥1 |
| Dyspnea (while resting or walking on flat surface), n | 6 (4.3%) | 8 (2.9%) | 5 (10.4%) | 2 (16.7%) |
| Angina pectoris (current or previous), n | 10 (7%) | 26 (9.5%) | 9 (20%) | 0 (0%) |
| Diabetes (current or previous), n | 8 (5.6%) | 13 (4.6%) | 5 (10.6%) | 2 (18.2%) |
| Chest pain (current or previous, while walking up hills or stairs, or fast or normal pace on level ground), n | 20 (13.3%) | 37 (13.1%) | 5 (10.2%) | 2 (16.7%) |
| High blood pressure (current or previous), n | 72 (49%) | 114 (40.7%) | 28 (56%) | 8 (66.7%) |

| | | | | |
|---|---|---|---|---|
| **Heart rate, mean** | **61 (1.6)** | **61 (1.2)** | **62 (3.2)** | **71.8 (6.2)** |
| **BMI [kg/m^2], mean** | **26.4 (0.7)** | **26.2 (0.5)** | **26.9 (1.2)** | **28 (2.7)** |
| **Smoker (current or previous), n** | **87 (58.4%)** | **165 (57.5%)** | **30 (61.2%)** | **8 (61.5%)** |
| **Missing data** | | | | |
| | **AR≥3** | **MR≥3** | **AS≥1** | **MS≥1** |
| **Dyspnea missing, n** | **9 (6.0%)** | **16 (5.5%)** | **3 (5.9%)** | **1 (7.7%)** |
| **Angina missing, n** | **8 (5.3%)** | **17 (5.8%)** | **6 (11.8%)** | **4 (30.8%)** |
| **Diabetes, missing n** | **6 (4%)** | **12 (4.1%)** | **4 (7.8%)** | **2 (15.4%)** |
| **Chest pain, missing n** | **0 (0%)** | **10 (3.4%)** | **2 (3.9%)** | **1 (7.7%)** |
| **High blood pressure, missing n** | **3 (2%)** | **12 (4.1%)** | **1 (2%)** | **1 (7.7%)** |
| **Heart rate, missing n** | **5 (3.3%)** | **25 (8.6%)** | **5 (9.8%)** | **1 (7.7%)** |
| **Smoker missing, missing n** | **1 (0.7%)** | **5 (1.7%)** | **2 (3.9%)** | **0 (0%)** |

**Table S1. Summary statistics for study population and dataset variables.**
The table provides an overview of VHD-prevalence, and summary statistics on sex, age, clinical variables, and prevalence of missing data (percentages of the latter 3 subtables show prevalence of the row-variable within the VHD-subgroup of the column). VHD = ventricular heart disease. AR = Aortic regurgitation. MR = mitral regurgitation. AS = aortic stenosis. MS = mitral stenosis. SD = standard deviation. BMI = body mass index.

* Chest pain variable represents chest pain experienced when walking up hills or stairs, or walking fast on level ground, or when walking at normal pace at level ground.

| subgroup | AR | | MR | | AS | | MS | |
|---|---|---|---|---|---|---|---|---|
| | n (%) | RR (CI) | n (%) | RR (CI) | n (%) | RR (CI) | n (%) | RR (CI) |
| VHD grade ≥1 | 129 (28.0%) | 1.73 (1.46-2.06) | 224 (20.8%) | 1.17 (1.06-1.29) | 47 (92.2%) | 52.4 (18.99-145) | 12 (92.3%) | 53.52 (6.98-410) |
| VHD grade ≥2 | 77 (30.1%) | 1.92 (1.5-2.45) | 171 (21.2%) | 1.2 (1.06-1.37) | 31 (96.9%) | 138.3 (18.93-1010) | 3 (100%) | - |
| VHD grade ≥3 | 53 (35.3%) | 2.44 (1.78-3.34) | 75 (25.7%) | 1.54 (1.21-1.96) | 6 (100%) | - | 1 (100%) | - |
| VHD grade ≥4 | 30 (45.5%) | 3.72 (2.32-5.96) | 22 (31.9%) | 2.09 (1.27-3.42) | - | - | - | - |

**Table S2. Statistical relationship between VHD and murmur grade.**
The table shows the number and prevalence of participants with (mean) murmur-grade≥1 within the subgroup defined by the indicated VHD type and VHD-grade threshold. The table also shows risk-ratios (95% CI in parenthesis) associated with presence of murmur, comparing VHD risk for those with and without murmur. Some risk-ratios could not be computed due to there being no cases in the positive class for which there was absence of murmur, resulting in division by zero. RR = risk ratio. AR = Aortic regurgitation. MR = mitral regurgitation. VHD = ventricular heart disease. AS = aortic stenosis. MS = mitral stenosis. CI = confidence interval.

| | pos. 1 (aortic) | pos. 2 (pulmonic) | pos. 3 (tricuspid) | pos. 4 (mitral) | all pos. combined |
|---|---|---|---|---|---|
| p-value | 0.4715 | 0.0195* | 0.0488* | 0.0237* | 0.0131* |

**Table S3. Performance comparison: regression vs classification**
The table shows the results from a comparison between a network trained to predict murmur grade≥2, and a network trained to predict murmur grade as a continuous variable. The p-values correspond to the hypothesis that the AUC of the regression model is higher than the classification model trained on a binary output (murmur grade≥2). Each column shows the result when predictions are made on audio from the index position. For the rightmost column, the algorithms made predictions on audio from all 4 auscultation positions, and the combined AUC was considered.

#### Comparison Between Continuous and Binary Labels

When training a murmur detection algorithm, we may model the problem as either a regression problem or as a classification problem. In the former approach, we model the annotated murmur grade as a continuous variable, and in the latter we model it as a binary variable, with prediction classes being "murmur" and "no murmur". In the reduction of the data to binary form there is potentially a loss of information. For instance, if one recording is annotated as grade 1 and another as grade 0, it is appropriate for these two samples to assign different loss values to e.g. a prediction of grade 2. If we instead used binary labels and a cutoff for the positive class of e.g. grade 2, then the distinction between the labels would be lost, and a prediction of grade 2 (or a corresponding activation value) would receive the same loss in both cases. We suspect therefore that the regression approach might produce a better model, as it plausably uses the training data more efficiently. Formally, our hypothesis is that the regression model will predict murmur grade≥2 more accurately than a classification model trained directly to predict such cases, with accuracy measured by the AUC.

We collect comparison data by performing 8-fold cross validation, using data from all positions jointly during training, with CV splits being based on participants ID to ensure independence between each training and validation set pair. For each fold, we compare AUC values, after which we use the paired t-test (two sided) to test for significant performance difference. We compare AUC for each position separately, as well as for all positions jointly (in each CV split, the 4 validation sets are joined into a single set for which the AUC is computed). In order to make the comparison as objective as possible, we use an automatic rule for training stoppage; training is stopped when the value of the loss function has failed to improve over 5 consecutive epochs (one check per epoch), with failure to improve being defined as failure to improve upon the preceding 4 values.

##### Result

The regression model significantly outperformed the classification model. Testing the hypothesis that the regression network predicted murmur grade≥2 with a higher AUC than the classification network, the p-value obtained was 0.013 (**Table S3**). In this comparison, predictions for all recordings (positions 1-4) were used, with the success or failure of each prediction being treated as independent outcomes. Significant improvements were also seen for murmur prediction in positions 2, 3, and 4, but not in position 1 where the performance was not significantly different.

The aortic position had approximately twice as many cases of murmur-grade≥2 as any of the other positions. It is possible that, due to making less efficient use of the data, the classification network is more prone to overtraining than the regression network, and therefore it overfits to the aortic position, which would explain why we see significant outperformance in all other positions. In any case, it seems that modeling murmur grade as a continuous variable is likely a more efficient way of training a murmur detection algorithm than splitting the label data into two classes and modeling as a classification problem.

#### Modified Version of Springers Segmentation Algorithm (patent pending)

The segmentation algorithm proposed by Springer et. al. in 2015 is probabilistic heart sound (HS) segmentation algorithm that estimates the cardiac states in a HS-signal with a Hidden Markov Model. It attempts to estimate the most likely sequence of states, with possible states defined as S1, systole, S2, and diastole. To this day it remains a competitive and popular segmentation algorithm and benchmark method.

Prior to maximum likelihood based assignement of cardiac states, Springer's segmentation algorithm estimates the average heart rate (HR) and systole duration of the audio signal via the autocorrelation function (ACF) of the (pre-processed) signals homomorphic envelope. Accurate estimation of these parameters is essential for the succsess in subsequent segmentation. To identify the HR-peak (the peak corresponding to the HR) the maximum over the interval $I_{search} = [0.5s, 2s]$ is identified, and the time lag corresponding to this peak is used to estimate the heart rate.

We have made modifications to this approach which accounts for situations in which peaks that occur at integer multiples of the time lags of the HR-peak are larger than the HR peak (**Figure S5** shows an example of this occuring), which would otherwise result in the heart rate estimate being off by an integer multiple. We call this Modification 1.

Furthermore, we have implemented a method for jointly using audio recordings from all four standard auscultation positions for a more robust estimation of the heart rates. The general idea is to provide each position the oppertunity to use the HR-estimate of another position with cleaner audio based on suitable quality metrics as well as other metrics that are relevant for detecting erroneous estimates. We refer to this method as Modification 2.

### Modification 1

The 3 highest peaks of the ACF in $I_{HR}$ are identified, with coordinates $(x_i, y_i)$ which we denote the *candidate peaks*. After identifying the *maximum peak*, with coordinates denoted $(x^*, y^*)$, tolerance windows in the x and y directions are calculated, $I_x = [0.85 \cdot 1/2 \cdot x^*, 1.15 \cdot 1/2 \cdot x^*] = [0.425 \cdot x^*, 0.575 \cdot x^*]$ and $I_y = [0.6 \cdot y^*, \text{inf}]$, which are used to determine if the maximum peak is a repeat of a smaller identified peak. If one of the smaller peaks $(x_i, y_i)$ lies within the set $I_x \times I_y$, then it is concluded that the $x^*$ is a multiple of $x_i$, and that $(x_i, y_i)$ is the true HR-peak. **Figure S5** shows a graphic illustration of how the modification identifies the true HR peak.

### Modification 2

The recordings of the different positions often varied considerably in terms of how pronounced and regular S1 and S2 were, and consequently how reliably the heart rate could be estimated from the ACF. As the average heart rate typically varies little between the recordings of an individual, we wondered if performance could be improved by performing a joint heart rate estimation where recordings with a weak signal can utilize information from recordings with a more distinct and regular pattern. We implemented a simple version of this idea, along the following lines:

1. Estimate the average heart rate in each position.
2. Calculate a *confidence score* $c_i$ for each position, based on features of the heart rate peak and the ACF that reflects the likelihood that the heart rate estimate is reliable.
3. Allow a position $i$ to discard its estimate in favour of the estimate of a more confident *neighbour* if its confidence score $c_i$ is low, and if its estimate deviates strongly from the estimates of the other positions.

The confidence score $c_i$ of a position $i$ is calculated based on three properties of the corresponding ACF:

1. Periodicity $S_{periodicity}$, as measured by how well it can be approximated with a periodic function.
2. Height of the HR-peak relative to the other peaks in $I_{HR}$, $S_{rel.\ height}$.
3. Peak prominence, $S_{prominence}$, wich measures how pronounced the peak is by its height relative to the adjacent valleys.

##### Periodicity

As the second half of the ACF provides little information, it is discarded. For estimation of periodicity, we first capture the slow changing trend of the ACF with a smoothing function (gaussian kernel, using the matlab function 'smoothedata') using a window length of 4.57s, after which this trend, $M_{trend}$, is subtracted from the ACF to form the de-meaned ACF $r_0$. We then compute the discrete cosine transform of $r_0$, and discard all but the four largest coefficients, thus obtaining a periodic function $G_{periodic}$ which approximates $r_0$. Finally, we score periodicity based on how well $G_{periodic}$ approximates $r_0$ relative to a benchmark fit provided by the smoothing function $M_{trend}$, with goodness of fit measured in by the root-mean-squared-error (RMSE) of the difference:

$$S_{periodicity} = 1 - \text{RMSE}(G_{periodic} - r_0)/\text{RMSE}(M_{trend} - r_0)$$

##### Peak Prominence and Relative Peak Height

Peak prominence $S_{prominence}$ is a local measure of peak prominence, and reflects the distinctiveness of a peak relative to its local neighbourhood. This is achieved by locating the adjacent valleys (one on the left, and one on the right, see the MATLAB documemtation page for details) and computes the height of the peak relative to the highest of the two neighbouring valleys.

Relative peak height $S_{rel.height}$ measures the prominence of the HR peak relative to the height of the second highest peak in $I_{HR}$. It measures peak prominence in a more global sense by calculating the ratio $y_{HR}/y'$, where $y_{HR}$ is the height of the HR-peak, and $y'$ is the height of the tallest peak within $I_{HR}$ that is not the HR peak.

##### Confidence Score and Decision Rule

The confidence scores $c_i$ are computed according to the formula:

$$c_i = S_{prominence} \cdot S_{periodicity} \cdot (1.5 \cdot \text{sigmoid}(S_{rel.\ height} - 1.4) + 0.9);$$

Let $h_i$ denote the HR-estimate of position $i$. Define

$$\bar{h} = \sum_{i=1}^{4} \left( h_i \cdot \frac{c_i}{\sum_{j=1}^{4} c_j} \right)$$

which is a weighted average of the HR-estimates, with weights determined by the confidence scores. Furthermore, define $d_i = |h_i - \bar{h}|$, $c^* = \max(\{c_i : 1 \leq i \leq 4\}$ (the highest confidence score, with corresponding index $i^*$), and

$$\sigma_{-i} = 1/2 \cdot \sqrt{\sum_{j \neq i} (h_j - \bar{h}_{-i})^2},$$

i.e. the standard deviation of the estimates when excluding position $i$.
We want high deviation $d_i$ from the average estimate $\bar{h}$ and high agreement amongst the other positions (as measured by $1/\sigma_{-i}$) to increase the tendency for position $i$ to discard its own estimate and replace it with the most confident estimate $h^*$. We model this by defining the decision variable

$$\tilde{d_i} = \left( \frac{d_i}{1 + 1.4 \cdot \text{sigmoid}(0.25 \cdot \sigma_{-i})} \right)^{1.5}.$$

Similarly, we want high relative confidence $c_i/c^*$ and absolute confidence $c_i$ to decrease the tendency for position $i$ to discard its estimate, which we model with the decision variable $\tilde{c}_i$:

$$\tilde{c}_i = 0.22 \cdot (c_i/c^*)^{1.5} + 0.08 \cdot c_i.$$

$\tilde{c}_i$ and $\tilde{d}_i$ are combined to form the final decision variable $D_i$:

$$D_i = \tilde{c}_i - 0.09 \cdot \tilde{d}_i$$

The desicion rule is as follows: if $D_i < 0.78$, then position $i$ discards its HR-estimate $h_i$ and replaces it with $h^*$, otherwise it keeps its own estimate. In the case that all positions have low confidence scores (below a fixed threshold), the median of the HR-estimates is used for all positions.

Note that the numerical values presented here are the tuning values that we used in the present study.
